# Previously defined variants of uncertain significance may play an important role in epilepsy and interactions between certain variants may become pathogenic

**DOI:** 10.1101/2023.08.10.23293930

**Authors:** Yara Hussein, Hila Weisblum-Neuman, Bruria Ben-Zeev, Shani Stern

## Abstract

**Objective:** Epilepsy is a chronic neurological disorder related to various etiologies and the prevalence of active epilepsy is estimated to be between 4-10 per 1000 individuals having a significant role of genetic mutations. Next-Generation Sequencing (NGS) panels are utilized for genetic testing, still, a substantial proportion of the results remain uncertain and are not considered directly causative of epilepsy. This study aimed to reevaluate pediatric patients diagnosed with epilepsy who underwent genetic investigation using NGS panels, focusing on inconclusive variant findings or multiple Variants of Uncertain Significance (VUSs).

**Methods:** A subgroup of pediatric patients aged 0-25 years, diagnosed with epilepsy, who underwent genetic investigation with an NGS epilepsy panel at the Child Neurology Unit, The Edmond and Lily Safra Children’s Hospital, Sheba Medical Center, between 2018-2022 through Invitae, was reevaluated. Patients with inconclusive variant findings or multiple VUSs in their test results were included. Genetic data were analyzed, focusing on identifying potentially pathogenic variants and frequent genetic combinations.

**Results:** Two unrelated potentially pathogenic variants were identified in the SCN9A and QARS1 genes. A frequent genetic combination, RANBP2&RYR3, was also observed among other combinations. The RANBP2 gene consistently co-occurred with RYR3 variants in uncertain results, suggesting potential pathogenicity. Analysis of unaffected parents’ data revealed certain combinations inherited from different parents, suggesting specific gene combinations as potential risk factors for the disease.

**Significance:** This study highlights the importance of reevaluating genetic data from pediatric epilepsy patients with inconclusive variant findings or multiple VUSs. Identification of potentially pathogenic variants and frequent genetic combinations, such as RANBP2&RYR3, could aid in understanding the genetic basis of epilepsy and identifying potential hotspots.

**Plain Language Summary:** We have performed a retrospective analysis on a subpopulation of pediatric patients diagnosed with epilepsy, we have found that specific genetic variants were repeatable indicating their potential pathogenicity to the disease.

**Key points:**

- 60% of pediatric patients undergoing genetic testing receive an uncertain result emphasizing the complexity of genetic interpretation in epilepsy diagnostics.
- Pathogenic variants in genes like SCN1A were common, underlining the importance of targeted gene sequencing.
- Variants in genes like SCN9A and QARS1, currently classified as VUSs, showed consistent presence in epilepsy patients, indicating potential pathogenicity.
- Specific genetic combinations, such as RANBP2&RYR3, were frequently observed among uncertain results, suggesting potential pathogenicity.

## Introduction

Epilepsy is a chronic neurological ailment distinguished by a persistent susceptibility to generate seizures and incur subsequent neurobiological, cognitive, psychological, and social ramifications arising from recurrent seizure occurrences^1,2^.

According to the World Health Organization (WHO), the prevalence of active epilepsy is estimated to be between 4-10 per 1000 individuals in the general population; On a global scale, approximately 5 million people are diagnosed with epilepsy yearly^3^ while the majority of epilepsy is diagnosed during pediatric years in which around 1 in 150 children is diagnosed, with the highest incidence rate occurring in infancy ^4^.

Epilepsy can be attributed to various causes but the genetic component is strong; Over 50% of epilepsy cases have a genetic component^5,6^. The literature has described over 500 genes associated with epilepsy, suggesting their potential contribution to the development of the condition^7^. Various genes were found to be highly associated with epilepsy including genes coding for sodium channel subunits, GABA receptor subunits, and others^8,9^. Electrophysiological studies on patient-derived cortical and hippocampal neurons with mutations associated with epilepsy show a neuronal hyperexcitability pattern ^11–15^ and a reduction in GABA-positive neurons^11,12^.

The diagnosis of epilepsy relies primarily on a detailed patient history and neurological examination, while EEGs and neuroimaging like MRI and CT scans serve as critical adjuncts. EEGs, enhanced by state-dependent recordings and activation procedures, help identify epileptiform abnormalities, though they may require intracranial monitoring for elusive cases. Additionally, metabolic, and genetic evaluations are tailored to the seizure type and suspected syndrome, with an increasing role for comprehensive genetic testing in identifying etiologies of epileptic encephalopathies^16^.

Traditional genetic diagnostic tools including Whole Exome Sequencing (WES) employ sequencing on the protein-coding regions of a genome through oligonucleotide probes for targeted enrichment of the exome, capturing all coding regions within the genome for sequencing^17^. Lately, Next-Generation Sequencing (NGS) panels, a sequencing technology, is becoming a powerful diagnostic tool used for epilepsy. This method allows for the high-throughput analysis of DNA and RNA, generating sequencing data rapidly and cost-effectively compared to traditional methods. NGS panels involve several steps, including DNA fragmentation, library preparation, sequencing by synthesis, and data analysis, enabling the simultaneous sequencing of multiple DNA fragments^18^. Based on this method, custom panels, in which disease-relevant genomic regions are sequenced, were designed to focus on particular areas of the genome that are of interest for specific research questions or clinical applications balancing specificity with cost-efficiency^19,20^.

The yield of positive molecular tests is relatively low, about 15%-25% are categorized as pathogenic or likely pathogenic^10,21^; On the other hand, most of the variants found in these epilepsy panels are categorized as variants of uncertain significance (VUS)^10^, posing a substantial challenge in contemporary genetic variation screening approaches and genetic counseling.

The aim of our work is to reevaluate a subgroup of tested individuals with inconclusive variant findings, or more than one VUS in their panel results, assuming that several rare variants in genetic hotspots can lead to epilepsy manifestation, yet not categorized as pathogenic.

## Materials & Methods

A retrospective analysis was performed on a cohort of a single tertiary center for pediatric patients at the Child Neurology Unit, The Edmond and Lily Safra Children’s Hospital, Sheba Medical Center, and selected unaffected biological parents, that were genetically tested through the Invitae epilepsy panel as part of clinical assessments between the years 2018-2022, after obtaining formal approval from the Institutional Review Board (IRB) of Sheba Medical Center, ensuring full compliance with established guidelines and standards for research involving human subjects.

### Invitae Epilepsy testing

Pediatric patients at the Child Neurology Unit, The Edmond and Lily Safra Children’s Hospital, Sheba Medical Center were genetically tested through the Invitae epilepsy panel as part of clinical assessments between the years 2018-2022. Invitae epilepsy tests utilize a targeted gene panel based on NGS technology in which genomic DNA was extracted from blood or saliva samples and enriched for specific regions of interest using a hybridization-based protocol, followed by sequencing using Illumina technology. The sequencing depth for all targeted regions was set at a minimum of 50 times, and the resulting reads were aligned to the GRCh37 reference sequence. The analysis focused on the coding sequence of the indicated transcripts, including a 10-base pair flanking intronic sequence as well as specific genomic regions known to be causative of disease. The aim is to identify single-nucleotide variants (SNVs), short and long indels, exon-level deletions/duplications, and rare structural rearrangements that disrupt coding sequences performed by Invitae. The panel targets genes associated with both syndromic and nonsyndromic causes of epilepsy, generally 187 genes, as depicted in the latest version of Invitae until 2022 with various add-ons, providing a comprehensive assessment of the genetic factors underlying the condition.

### Variants Classification

Observed variants are classified as pathogenic (directly contribute to the development of a disease), likely pathogenic (likelihood of causing a known genetic condition), Benign (not known to cause genetic conditions, but can alter the protein products or change the gene’s expression), likely benign (is not expected to lead to a genetic condition, but the scientific evidence is not as strong as for the variants that are classified benign) or variant of uncertain significance (VUS) which is a variation in DNA that has an uncertain or unknown impact on health as provided by Invitae. These classifications are based on Sherloc-Invitae’s variant classification algorithm, based on the initial American College of Medical Genetics and Genomics/Association for Molecular Pathology (ACMG/AMP) classification framework, and represent the industry standard among clinical genetic testing laboratories^22^. In order to minimize uncertainty in genetic testing, follow-up testing for selected unaffected biological parents of patients previously tested was done.

### Observed Variants analysis

The electronic tests of 156 probands and 74 unaffected biological parents performed in the years 2018-2022 in the Edmond and Lily Safra Children’s Hospital, Sheba Medical Center, were used and data were extracted using custom-written MATLAB scripts (R2023a, Mathworks) and anonymously analyzed (included and excluded data are shown in supplementary Fig S1a).

Tests were grouped into 3 groups, Positive tests (in which there was a detection of at least one pathogenic/likely pathogenic variant that directly contributes to the emergence of epilepsy), Negative tests (no detected variant in the selected genes or only benign or likely benign detected variants) and Uncertain tests (at least one detected VUS). For each, the total number of detected variants was counted, and the most definitive leaders were found and compared to their population incidence based on the gnomAD browser (https://gnomad.broadinstitute.org/). Protein network path analysis was performed to find significant pathways against the Kyoto Encyclopedia of Genes and Genomes (KEGG) and Gene Ontology (GO) databases for observed pathogenic and uncertain variants separately. The Network Analyst web application was used to create a graphical representation of Protein network path analysis (https://www.networkanalyst.ca).

In the collective findings, the most prevalent combinations of VUS were identified across each subgroup. An identical analysis was conducted on probands who possessed data for both unaffected biological parents, aiming to identify notable combinations of genetic variants, genetic correlations were calculated using Correlation AnalyzeR (https://gccri.bishop-lab.uthscsa.edu/shiny/correlation-analyzer/).

## Results

156 pediatric patients at the Child Neurology Unit, The Edmond and Lily Safra Children’s Hospital, Sheba Medical Center, underwent genetic testing using the Invitae epilepsy panel (Blood samples = 94%). Approximately half were females (53.2%) and the mean age at test requisition was 7.6 ± 5.3 years (range: 0-25 years). The epilepsy panel is occasionally supplemented with additional panels or add-ons; On average, a total of 227.4 + 59.7 genes were tested (with the main panel consisting of 187 genes, as depicted in the latest version of Invitae until 2022, as shown in supplementary figure S1b). Genetic panel tests were performed for various indications, including Refractory epilepsies, Infantile spasms, Self-limited childhood focal epilepsies accompanied by Electrical status epilepticus in sleep (ESES), focal, and generalized epilepsies. Most of the patients had Refractory epilepsy, while the minority responded to monotherapy. The population was also versatile in her characteristics regarding other neurologic manifestations: Developmental delay, autistic spectrum disorder (ASD), Intellectual disability, mood disorder, anxiety, learning difficulties, speech difficulties, behavioral difficulties, attention deficit and hyperactivity disorder (ADHD), ticks, headaches, weakness episodes dizziness and tinnitus.

A total of 18% of the probands who underwent testing received a negative result, indicating no detectable genetic variants related to epilepsy in the performed panel, 22% of the probands obtained a positive result, indicating the presence of at least one pathogenic variant associated with epilepsy. The majority, constituting 60% of the probands, received an uncertain result (see methods) as shown in Figure 1a and supplementary Figure S1c.

**Figure 1.**
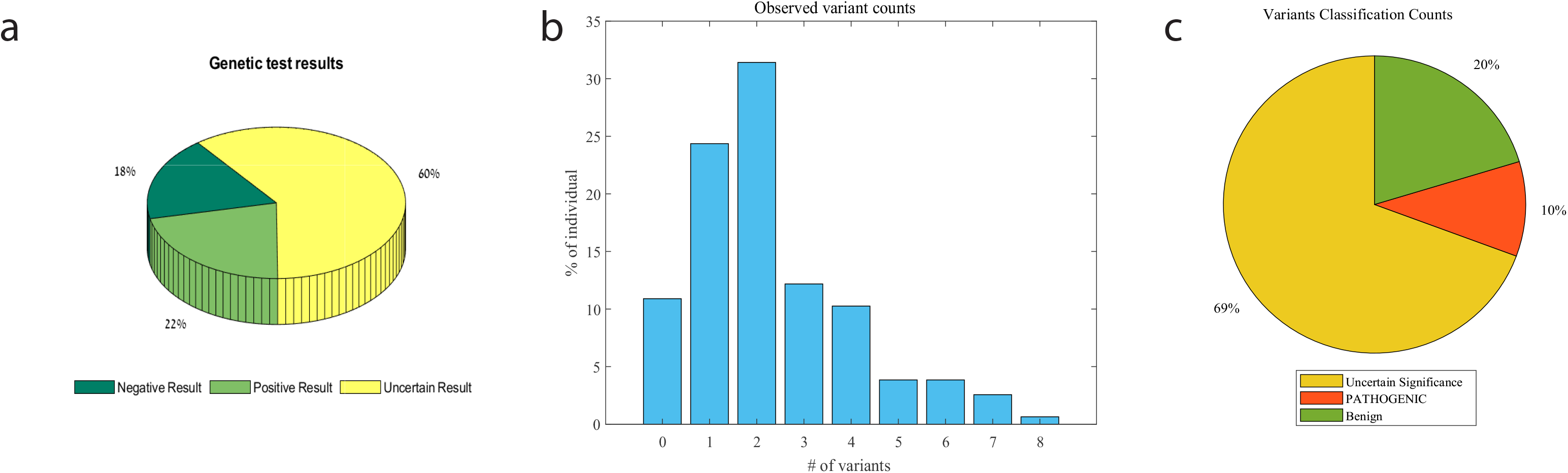
A graphical description of the probands data. **(a)** Test outcomes categorized according to the observed genetic variants. **(b)** Observed genetic variants in the targeted genes on the NGS Invitae panel ranged from zero to 8 per individual. **(c)** A classification of the observed variants.

On average, there were 2.3 + 1.7 observed genetic variants across the probands. Notably, 11% of the individuals showed no observed variants in the targeted genes at all, while almost 65% exhibited 2 or more observed variants, as shown in Figure 1b. Among all the observed variants, 20% were classified as benign or likely benign, 10% were classified as pathogenic or likely pathogenic, and the majority, constituting 69%, were classified as variants of uncertain significance as shown in Figure 1c.

### Regulation of action potential and multiple neuronal developmental processes are dysregulated in child epilepsy

As previously mentioned, 22% of the tests were positive, out of these 15% exhibited the presence of two different pathogenic variants, while 85% indicated the presence of a single pathogenic variant contributing to the onset of epilepsy (Figure 2a). Among the pathogenic genes identified in the probands, the SCN1A gene-coding for the α-subunit of a neuronal voltage-gated sodium channel^23^ was detected in 11% of the positive results, followed by MECP2 and CDKL5 genes, each observed in 8% of the cases as shown in Figure 2b. Noteworthy the pathogenic c.1274_1277dup in the HEXA gene, the c.1385G>C variant in the PNKP gene, the c.649dup in the PRRT2 gene, and the c.671A>G variant in the GCHI gene, were observed twice each in the positive tests (Figure 2c). Notably, mutations in the HEXA and PNKP genes are associated with autosomal recessive conditions, while mutations in the PRRT2 and GCHI genes are associated with autosomal dominant conditions suggesting their potential pathogenicity role in explaining the epileptic condition.

**Figure 2.**
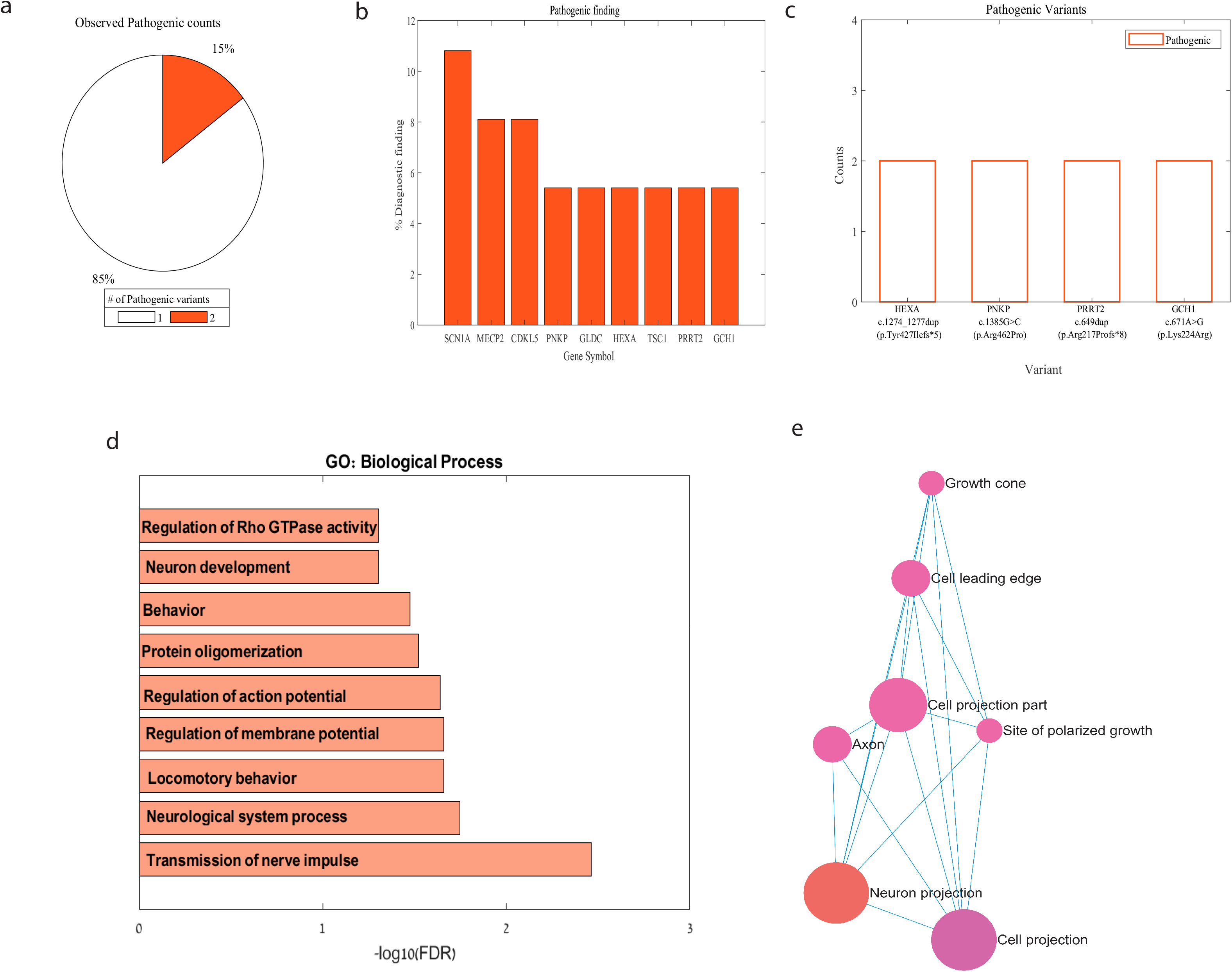
Diagnostic Results and genes with pathogenic findings. **(a)** Among individuals with positive results, the observed pathogenic variants in the targeted genes on the NGS Invitae panel ranged from 1 pathogenic variant responsible for the condition to two pathogenic variants per individual. **(b)** The pathogenic genes repeated more than three times in the positive proband data. **(c)** The pathogenic variants repeated more than once in the positive proband data. **(d)** Significant enrichment of affected GO biological process in the set of pathogenic genes (FDR < 0.05). **(e)** Significant enrichment of affected GO cellular components in the set of pathogenic genes (FDR < 0.05).

Based on the detected pathogenic genes in our tests, Cellular Components (CC) related to neuron projection, cell projection, axon, growth cone, and others showed significant GO: CC enrichment (FDR<0.05). In terms of biological processes (BP), the transmission of nerve impulses, neurological system processes, regulations for action and membrane potentials, and neuron development were enriched (FDR < 0.05), as shown in Figure 2d-e indicating several neuronal alterations^24^ contributing to the development of epilepsy.

### Two genetic alterations in the SCN9A and QARS1 genes, currently classified as VUS’s have a potential implication for pathogenicity in child epilepsy

For the uncertain results with at least one detected VUS, an average of 2 + 1.2 VUSs were present in the patients’ panels, out of which 45% showed the presence of a single VUS, while 55% exhibited two or more VUSs, as presented in Figure 3a. SCN5A, FASN, and RYR3 were the most frequently observed VUS genes as shown in Figures 3b and 3c. Furthermore, the c.2133G>C variant in the SCN9A gene and the c.316G>A variant in the QARS1 gene were observed three times each in the uncertain tests, along with other variants shown in Figure 3d. These variants appear to have a high penetrance in epilepsy patients compared to their presence in the general population based on the gnomAD database (p=3.76e-07) although classified to be with an uncertain significance. The clinical background of affected patients is shown in Supplementary Tables 1,2.

**Figure 3.**
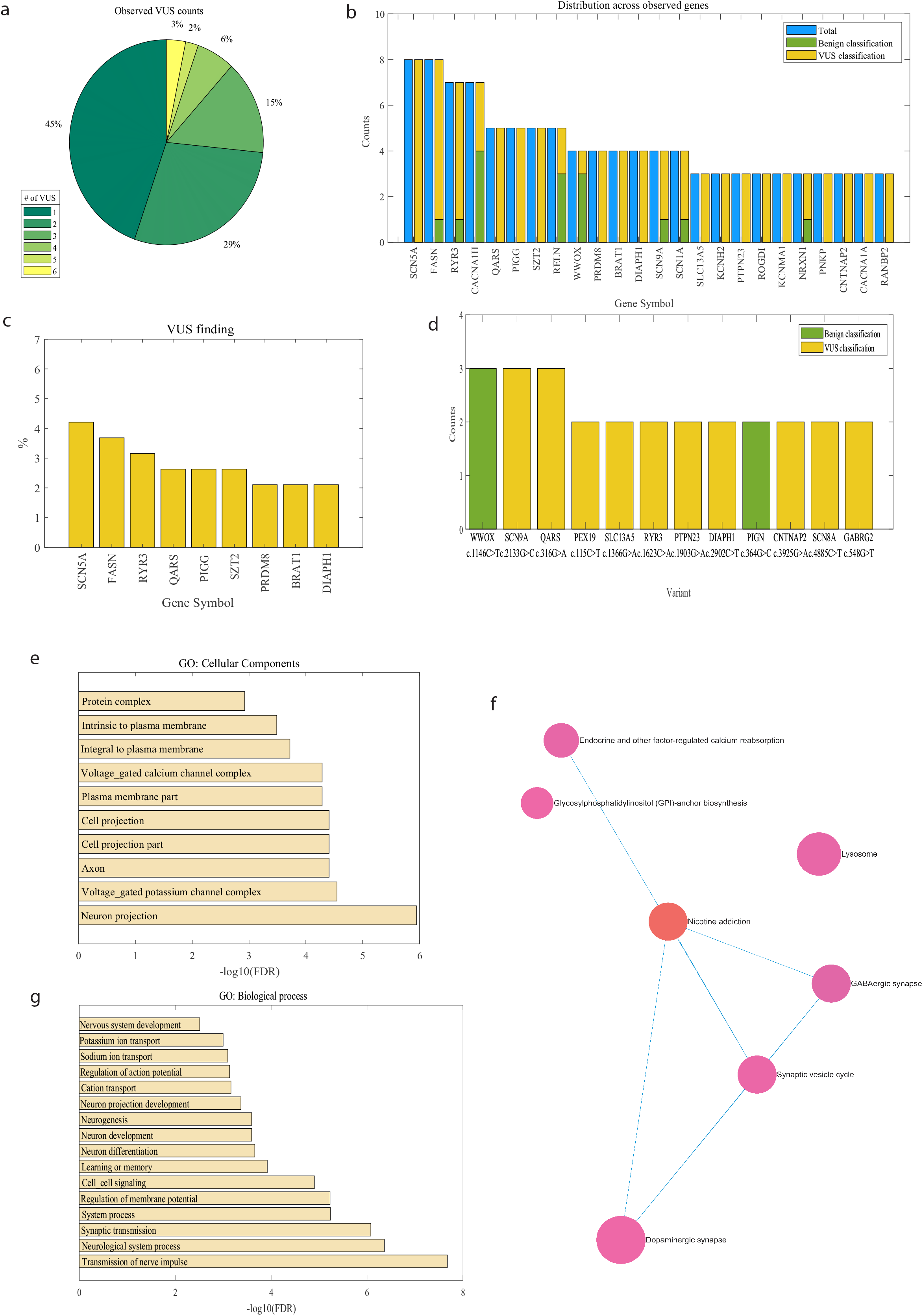
Uncertain Results and genes with uncertain findings. **(a)** Among individuals with uncertain results, the observed uncertain variants in the targeted genes on the NGS Invitae panel range from one to six variants per individual. **(b)** The frequent genes in the uncertain data. **(c)** The frequent genes with uncertain yield. **(d)** The uncertain Variant yield frequencies. **(e)** Significant enrichment of affected GO cellular components in the set of VUS genes (FDR < 0.05). **(f)** Significant enrichment of affected KEGG pathways in the set of pathogenic genes (FDR < 0.05). **(g)** Significant enrichment of affected GO biological process in the set of VUS genes (FDR < 0.05).

In terms of GO biological processes, the detected variants of uncertain significance showed enrichment in processes related to the transmission of nerve impulses, neurological system processes, regulations for action and membrane potentials, cell signaling, synaptic transmission, neuron development, and differentiation, among others. Furthermore, various neuronal components and channels such as voltage-gated calcium and potassium channels were enriched in terms of GO: CC (FDR < 0.05), (Figure 3e and Figure 3g). Notably, KEGG pathways associated with different types of synapses, including GABAergic and dopaminergic synapses and the synaptic vesicle cycle, showed significant enrichment (FDR < 0.05) as shown in Figure 3f.

### RANBP2 and RYR3 variant combination as a risk factor for child epilepsy

Upon analyzing the frequent combinations of VUSs in the overall results, it became evident that the RANBP2&RYR3 variant combination occurred most frequently; It was observed three times in our data. Additional combinations of VUS are shown in Figure 4a. Similarly, the RANBP2 & RYR3 combination remains the most prevalent when focusing solely on uncertain results, as shown in Figure 4b; Interestingly, all detected variants of the RANBP2 gene were designated as VUSs and coexisted in diverse combinations with the RYR3 variants. The clinical background of affected patients is shown in Supplementary Table 3.

**Figure 4.**
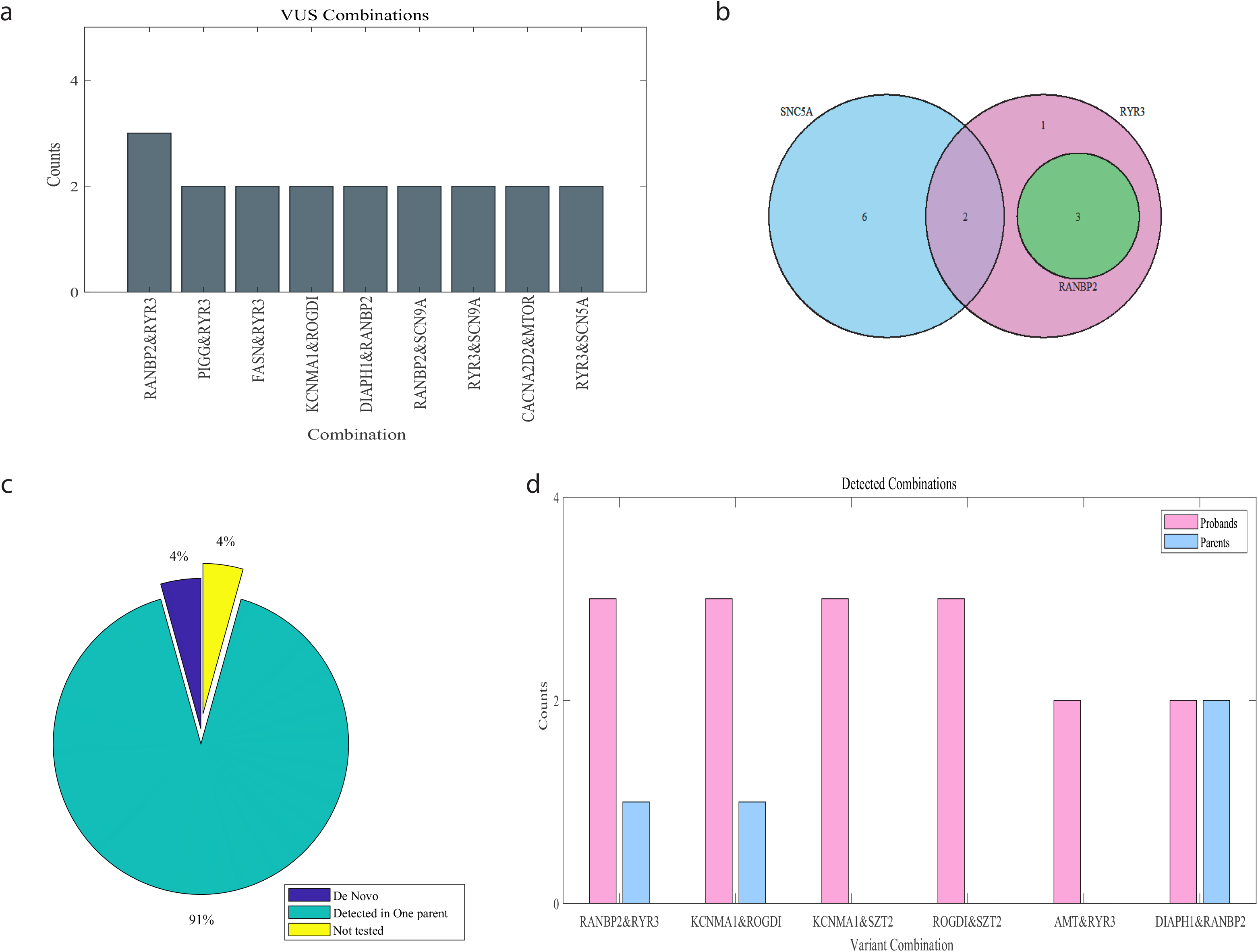
Genetic Combinations as a hotspot for child epilepsy. **(a)** Frequent genetic combinations among affected children. **(b)** Frequent genetic combinations among uncertain results in the SCN5A, Ryr3, and RANBP2 genes. **(c)** Distribution of De novo, Detected and not tested variants in parent’s data. **(d)** Frequent genetic combinations among individuals and unaffected biological parents

For a subgroup of selected probands (n=38) within the cohort, genetic testing was conducted on both unaffected biological parents (n=74) to assess the variants identified in their children. Remarkably, 91% of the observed proband variants were also found in one unaffected parent, while 4% of the variants were determined to be de novo, meaning they were not present in either parent, as illustrated in Figure 4c. To investigate potential combinations that may contribute to the risk of developing epilepsy, optional combinations in the parental data were analyzed. The combination of RANBP2 and RYR3 remained notably prominent among the observed variants in the proband data, as depicted in Figure 3d. We observed a single distinct inherited genetic combination, each originating from different parents, suggesting a potential genetic basis for the child’s epilepsy. Additionally, there was a single case where a parent shared the same genetic combination as their affected child, despite lacking an epilepsy diagnosis. Additionally, other combinations in probands data exhibited potential risk factors compared to their unaffected biological parents (p= 6.7471e-04).

## Discussion

In the past decade, significant progress has been made in identifying gene mutations associated with epilepsy and unraveling the molecular mechanisms that contribute to the clinical presentation of the disease. It is becoming increasingly evident that comprehending these mechanisms is crucial for selecting optimal treatment approaches for affected individuals. Consequently, genetic testing is done to determine whether these features are associated with a known genetic condition^25^.

Epilepsy, being a disorder with a notable genetic component^26^, has been linked to numerous genes known to play a direct role in its development. To facilitate the identification of potential pathogenic variants in these epilepsy-associated genes, NGS panels have been developed. These panels are designed to probe genes highly relevant to epilepsy, allowing for a targeted and comprehensive analysis of genetic variations^27^ such as Invitae’s epilepsy panel.

In this study, we analyzed genetic data from patients diagnosed with infantile and childhood epilepsy who had undergone Invitae’s tests. These samples were obtained either from blood or saliva, some with confirmed genetic alterations correlated with epilepsy, and some with no definite genetic condition. It is important to highlight that the tests’ results exhibited partial ambiguity, with approximately 60% of the tests (and 69% of the detected variants) showing significant uncertainty. One relatively unique characteristic of VUSs is that while the result itself may remain static, its meaning is often resolved over time, as more data are gathered^25^. This complex task is crucial for the comprehension of the underlying pathogenic elements, enabling the understanding of involved mechanisms as well as implementing the appropriate treatments leading to effectively managing the disorder.

Consistent with previously reported data^28^, we observed that the SCN1A gene was one of the leaders in the observed pathogenic variants resulting in epilepsy seizures; associated with Dravet syndrome (DS)^29^ and other epilepsy syndromes^30^.

Upon analyzing the VUSs reported, we observed that the c.2133G>C (p.Leu711Phe) variant in the SCN9A gene was repeated in 3% of our tests. This sequence change replaces leucine with phenylalanine, which are both neutral and non-polar amino acids at codon 711 of the SCN9A protein (rs772492538, gnomAD 0.02%, PolyPhen-2 prediction: Probably Damaging^35^) potentially altering the function of the Nav.17 sodium channel leading to signal disruptions^36^. Similarly, a repeated unrelated variant is the c.316G>A (p.Asp106Asn) variant in the QARS1 gene-inherited in an autosomal recessive pattern and associated with “Microcephaly, progressive, with seizures and cerebral and cerebellar atrophy” syndrome (OMIM #615760)^37^. We observed that this variant was repeated also in 3% of our tests. This sequence change replaces aspartic acid, which is acidic polar, with asparagine, which is a neutral polar amino acid, at codon 106 of the QARS protein (rs141983717, gnomAD 0.04%). The consistent observations of these variants in multiple unrelated patients indicate their potential pathogenicity, leading to their consideration as a causative variant responsible for epilepsy development. It is noteworthy that these specific variants have not been reported as associated with epilepsy yet.

Furthermore, based on the set of detected VUSs an observed dysregulation of several neurodevelopmental functions and GABAergic synapse enrichment as previously reported ^35^. A decreased activity of the inhibitory circuitry, mainly GABAergic, is thus likely to be a major factor contributing to seizure generation in affected patients^29^

Nearly half of the patients with an uncertain result showed the presence of two or more VUSs among their detected variants, highlighting the importance of precise interpretation regarding the clinical relevance of various variant combinations. Notably, the combination of RYR3&RANBP2 variants was observed multiple times in our data, standing out among all other possible combinations. Interestingly, the RANBP2 variants, when present as VUS, consistently co-occurred with a variant of RYR3, suggesting a potential pathogenic role for this combination in contributing to the manifestation of epilepsy.

The RANBP2 gene (on chromosome 2q11-13) encodes the nuclear pore component of RAN binding protein 2^38^ and is associated with acute necrotizing encephalopathy, an autosomal dominant ailment characterized by brain damage that usually follows an acute febrile disease^39^. The RYR3 gene is a ryanodine receptor predominantly expressed in the brain, and functions as a key regulator of calcium release from intracellular reservoirs. Its involvement in synaptic plasticity is well-established, and studies with RYR3 knockout mice have demonstrated compromised spatial learning abilities^40^. Following analyzing the detected variants from unaffected biological parents tests, it was observed that one RYR3 & Ranbp2 combination was inherited, with each variant originating from a different parent, suggesting that the combination of genes may be the cause of the child’s epilepsy. Notably, there was a single instance where one parent possessed the same combination as their affected child, despite not having a diagnosis. The increased prevalence of this genetic combination, relative to other potential combinations, suggests its potential role as a trigger or risk factor for disease development. However, it is crucial to note that the genetic correlation between these genes is quite low (Correlation AnalyzeR, r= 0.029; p=0.00805)^41^. Despite this, the observed pattern raises the possibility that specific gene combinations may influence disease manifestation or susceptibility. Further investigations are warranted to fully comprehend the implications and significance of these findings.

## Data Availability

All data produced in the present study are available upon reasonable request to the authors

## Acknowledgments

This work was supported by Zuckerman STEM Leadership Program and Israel Science Foundation (ISF) grants 1994/21 and 352/21.

## Author Contribution

SS designed, directed the study, and edited the manuscript, YH analyzed the data and wrote the manuscript, BBZ and HWN provided the electronic INVITAE tests and reviewed the manuscript.

## Data availability statement

Data supporting this study’s findings may be available upon reasonable request to the corresponding author.

## Ethical approval

This study was approved by the Institutional Review Board (IRB) of Sheba Medical Center ensuring full compliance with established guidelines and standards for research involving human subjects due to the retrospective approach.

## Figure Legends

**Figure S1.**
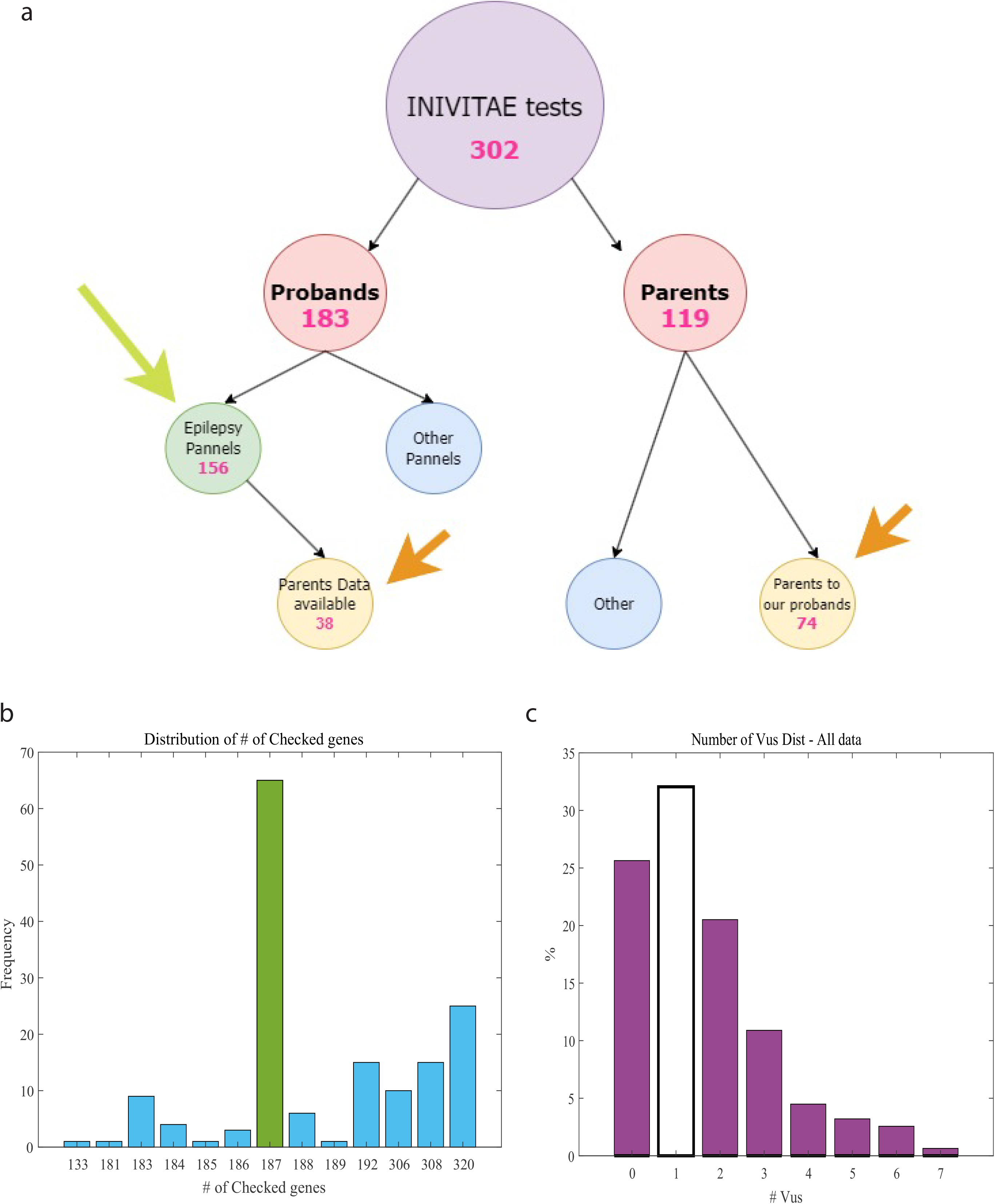
**(a)** Included and excluded data summary. **(b)** The distribution of targeted genes in the panels among individuals, the main epilepsy panel consists of 187 genes. **(c)** The observed uncertain variants in the targeted genes on the NGS Invitae panel range from zero to seven variants per individual.

## Notes

### Competing Interest Statement

The authors have declared no competing interest.

### Author Declarations

Ethics committee of The Sheba Fund for Health Services and Research gave ethical approval for this work

### Summary of Updates

title change and minor changes in the manuscript format

## References

1. Fisher, R. S. et al. Epileptic seizures and epilepsy: Definitions proposed by the International League Against Epilepsy (ILAE) and the International Bureau for Epilepsy (IBE). Epilepsia 46, 470–472 (2005).

2. Guerrini, R. Epilepsy in children. Lancet 367, 499–524 (2006).

3. World Health Organization. Epilepsy. https://www.who.int/news-room/fact-sheets/detail/epilepsy (2023).

4. Aaberg, K. M. et al. Incidence and prevalence of childhood epilepsy: A nationwide cohort study. Pediatrics 139, (2017).

5. Speed, D. et al. Describing the genetic architecture of epilepsy through heritability analysis. Brain 137, 2680–2689 (2014).

6. Pal, D. K., Pong, A. W. & Chung, W. K. Genetic evaluation and counseling for epilepsy. Nature Reviews Neurology vol. 6 445–453 Preprint at 10.1038/nrneurol.2010.92 (2010).

7. Weber, Y. G., Biskup, S., Helbig, K. L., Von Spiczak, S. & Lerche, H. The role of genetic testing in epilepsy diagnosis and management. Expert Rev Mol Diagn 17, 739–750 (2017).

8. Poduri, A. & Lowenstein, D. Epilepsy genetics—past, present, and future. Curr Opin Genet Dev 21, 325–332 (2011).

9. Wang, J. et al. Epilepsy-associated genes. Seizure vol. 44 11–20 Preprint at 10.1016/j.seizure.2016.11.030 (2017).

10. Lindy, A. S. et al. Diagnostic outcomes for genetic testing of 70 genes in 8565 patients with epilepsy and neurodevelopmental disorders. Epilepsia 59, 1062–1071 (2018).

11. Hussein, Y. et al. Early maturation and hyperexcitability is a shared phenotype of cortical neurons derived from different ASD-associated mutations. Transl Psychiatry 13, 246 (2023).

12. Brant, B. et al. IQSEC2 mutation associated with epilepsy, intellectual disability, and autism results in hyperexcitability of patient-derived neurons and deficient synaptic transmission. Mol Psychiatry 26, 7498–7508 (2021).

13. Repudi, S., Kustanovich, I., Abu-Swai, S., Stern, S. & Aqeilan, R. I. Neonatal neuronal WWOX gene therapy rescues Wwox null phenotypes . EMBO Mol Med 13, (2021).

14. Quraishi, I. H. et al. An epilepsy-associated KCNT1 mutation enhances excitability of human iPSC-derived neurons by increasing slack KNa currents. Journal of Neuroscience 39, 7438–7449 (2019).

15. Steinberg, D. J. et al. Modeling genetic epileptic encephalopathies using brain organoids. EMBO Mol Med 13, (2021).

16. Stafstrom, C. E. & Carmant, L. Seizures and epilepsy: An overview for neuroscientists. Cold Spring Harb Perspect Biol 7, 1–19 (2015).

17. Hodges, E. et al. Genome-wide in situ exon capture for selective resequencing. Nat Genet 39, 1522–1527 (2007).

18. Behjati, S. & Tarpey, P. S. What is next generation sequencing? Arch Dis Child Educ Pract Ed 98, 236–238 (2013).

19. LaDuca, H. et al. Exome sequencing covers >98% of mutations identified on targeted next generation sequencing panels. PLoS One 12, (2017).

20. Thermes, C. Ten years of next-generation sequencing technology. Trends in geneticsfZ: TIG vol. 30 418–426 Preprint at 10.1016/j.tig.2014.07.001 (2014).

21. Truty, R. et al. Possible precision medicine implications from genetic testing using combined detection of sequence and intragenic copy number variants in a large cohort with childhood epilepsy. Epilepsia Open 4, 397–408 (2019).

22. Nykamp, K. et al. Sherloc: A comprehensive refinement of the ACMG-AMP variant classification criteria. Genetics in Medicine 19, 1105–1117 (2017).

23. Escayg, A. et al. Mutations of SCN1A, encoding a neuronal sodium channel, in two families with GEFS+2. Nat Genet 24, 343–345 (2000).

24. Patel, D. C., Tewari, B. P., Chaunsali, L. & Sontheimer, H. Neuron–glia interactions in the pathophysiology of epilepsy. Nature Reviews Neuroscience vol. 20 282–297 Preprint at 10.1038/s41583-019-0126-4 (2019).

25. Hoffman-Andrews, L. The known unknown: The challenges of genetic variants of uncertain significance in clinical practice. J Law Biosci 4, 648–657 (2017).

26. Pal, D. K., Pong, A. W. & Chung, W. K. Genetic evaluation and counseling for epilepsy. Nature Reviews Neurology vol. 6 445–453 Preprint at 10.1038/nrneurol.2010.92 (2010).

27. Symonds, J. D. & McTague, A. Epilepsy and developmental disorders: Next generation sequencing in the clinic. European Journal of Paediatric Neurology vol. 24 15–23 Preprint at 10.1016/j.ejpn.2019.12.008 (2020).

28. Parrini, E. et al. Diagnostic Targeted Resequencing in 349 Patients with Drug-Resistant Pediatric Epilepsies Identifies Causative Mutations in 30 Different Genes. Hum Mutat 38, 216–225 (2017).

29. Dravet, C. The core Dravet syndrome phenotype. Epilepsia 52, 3–9 (2011).

30. Scheffer, I. E. & Nabbout, R. SCN1A-related phenotypes: Epilepsy and beyond. Epilepsia 60, S17–S24 (2019).

31. Catterall, W. A. Sodium channels, inherited epilepsy, and antiepileptic drugs. Annual Review of Pharmacology and Toxicology vol. 54 317–338 Preprint at 10.1146/annurev-pharmtox-011112-140232 (2014).

32. Escayg, A. & Goldin, A. L. Sodium channel SCN1A and epilepsy: Mutations and mechanisms. Epilepsia vol. 51 1650–1658 Preprint at 10.1111/j.1528-1167.2010.02640.x (2010).

33. Parisi, P. et al. Coexistence of epilepsy and Brugada syndrome in a family with SCN5A mutation. Epilepsy Res 105, 415–418 (2013).

34. Ma, H. et al. Mutations in the sodium channel genes SCN1A, SCN3A, and SCN9A in children with epilepsy with febrile seizures plus(EFS+). Seizure 88, 146–152 (2021).

35. Adzhubei, I. A. et al. A method and server for predicting damaging missense mutations. Nature Methods vol. 7 248–249 Preprint at 10.1038/nmeth0410-248 (2010).

36. Yang, Y. et al. Mutations in SCN9A, encoding a sodium channel alpha subunit, in patients with primary erythermalgia. J Med Genet 41, 171–174 (2004).

37. Zhang, X. et al. Mutations in QARS, encoding glutaminyl-trna synthetase, cause progressive microcephaly, cerebral-cerebellar atrophy, and intractable seizures. Am J Hum Genet 94, 547–558 (2014).

38. Neilson, D. E. et al. Infection-Triggered Familial or Recurrent Cases of Acute Necrotizing Encephalopathy Caused by Mutations in a Component of the Nuclear Pore, RANBP2. Am J Hum Genet 84, 44–51 (2009).

39. Hartley, M. et al. Acute Necrotizing Encephalopathy: 2 Case Reports on RANBP2 Mutation . Child Neurol Open 8, 2329048X2110307 (2021).

40. Balschun, D. et al. Deletion of the ryanodine receptor type 3 (RyR3) impairs forms of synaptic plasticity and spatial learning. EMBO Journal 18, 5264–5273 (1999).

41. Miller, H. E. & Bishop, A. J. R. Correlation AnalyzeR: functional predictions from gene co-expression correlations. BMC Bioinformatics 22, (2021).

